# Diagnostic accuracy of intraoperative frozen section in thyroid nodules with Bethesda III cytology: systematic review and meta-analysis

**DOI:** 10.64898/2026.07.25.26358934

**Authors:** José Luis Pardal-Refoyo, Ignacio Zapatero Sánchez

## Abstract

**Background:** Bethesda III thyroid nodules remain diagnostically indeterminate, and intraoperative frozen section is used selectively to support surgical decision-making. Its value in this specific cytological category is uncertain because a malignant result may be highly specific while non-malignant and non-definitive results may fail to exclude cancer.

**Objective:** To estimate the sensitivity and specificity of intraoperative frozen section for detecting malignancy in thyroid nodules with preoperative Bethesda III cytology.

**Methods:** The protocol was prospectively registered in PROSPERO (CRD420261416683). A systematic review was conducted in PubMed, Embase, Web of Science, Europe PMC, and the Cochrane Library. Studies were eligible when they reported a separable Bethesda III cohort, intraoperative frozen-section findings, and final histopathology. Frozen section was classified as positive only when malignancy was reported. Benign, suspicious, indeterminate, deferred, inconclusive, and follicular-pattern results were classified as non-malignant. Study-level 2 × 2 tables were synthesised with random-effects logit models. Because all studies reported zero false-positive results, a full bivariate model with freely estimated covariance was not identifiable; a pseudo-bivariate HSROC approximation was therefore used. QUADAS-2 was used for risk-of-bias assessment.

**Results:** Ten studies comprising 1,069 Bethesda III patients or nodules were included. The pooled sensitivity was 43.1% (95% CI, 21.3–67.9) and the pooled specificity was 98.8% (95% CI, 97.0–99.5). Sensitivity was highly heterogeneous (Q = 83.45, I^2^ >90%, τ^2^ = 2.15), whereas specificity showed negligible between-study variance. Excluding Mao 2023 reduced pooled sensitivity to 35.6% (95% CI, 23.8–49.5) and reduced sensitivity heterogeneity to moderate levels (Q = 12.44, I^2^ = 36%, τ^2^ = 0.25); specificity remained 98.7% (95% CI, 96.7–99.5). The approximate HSROC AUC was 0.89 including Mao 2023 and 0.87 excluding Mao 2023, but these values were driven largely by the uniformly high specificity.

**Conclusions:** Intraoperative frozen section in Bethesda III nodules has excellent specificity but limited and variable sensitivity. It is more suitable as a rule-in test than as a rule-out test. A non-malignant or non-definitive result should not be used alone to exclude malignancy or determine the extent of thyroid surgery.

## INTRODUCTION

The Bethesda System standardises reporting of thyroid fine-needle aspiration cytology. Bethesda category III includes atypia of undetermined significance and follicular lesion of undetermined significance, diagnoses that do not permit reliable classification as either benign or definitively neoplastic [1]. Subsequent management is individualised using clinical and ultrasound findings, repeat aspiration, molecular testing when available, surveillance, and diagnostic surgery [2, 3].

Intraoperative frozen section may identify unequivocal malignancy and permit completion of surgery during the same anaesthetic episode. However, the architectural and capsular findings needed to classify many follicular-pattern lesions may not be reliably assessed in frozen tissue. The potential value of frozen section therefore depends on a highly reliable positive result and on whether a negative result provides enough information to change management [4].

Diagnostic test accuracy reviews should synthesise sensitivity and specificity jointly, examine heterogeneity, and distinguish bias arising from patient selection, the index test, the reference standard, and patient flow [5, 6]. The present study followed the registered protocol and PRISMA-DTA principles [7, 8]. The objective was to estimate the diagnostic performance of intraoperative frozen section specifically in Bethesda III thyroid nodules and to examine the robustness of the findings through risk-of-bias assessment, leave-one-out analysis, and exclusion of an influential study.

## METHODS

### Review design and eligibility

The protocol was prospectively registered in PROSPERO (CRD420261416683).

This systematic review and meta-analysis evaluated diagnostic accuracy studies of intraoperative frozen section in Bethesda III thyroid nodules. The population comprised patients undergoing thyroid surgery for a nodule classified as Bethesda III or AUS/FLUS. The index test was intraoperative frozen section, intraoperative consultation, or an equivalent intraoperative pathological diagnosis. Final postoperative histopathology was the reference standard, and the target condition was thyroid malignancy.

Original human studies were eligible when the Bethesda III subgroup was separable and the published information allowed extraction or unequivocal reconstruction of true positives, false positives, false negatives, and true negatives. Mixed Bethesda cohorts were excluded when category III could not be separated. Reviews, editorials, non-human studies, and reports without usable diagnostic data were excluded.

### Information sources and study selection

Searches were conducted in PubMed, Embase, Web of Science, Europe PMC, and the Cochrane Library on July 7, 2026. No date, language, or study-design restrictions were documented. The full search strategies and database yields are provided in the accompanying search-strategy supplement.

The recorded selection process identified 221 records. After removal of 81 duplicates, 140 unique records were screened. Thirty-four reports were assessed in full text and 10 studies were included in the qualitative and quantitative synthesis. A more detailed numerical breakdown of full-text exclusion reasons was not available; therefore, the manuscript reports only the documented aggregate categories.

### Index-test classification and data extraction

Frozen section was classified as positive only when malignancy was reported. All other intraoperative categories were classified as non-malignant for the primary analysis, including benign, probably benign, suspicious, indeterminate, deferred, inconclusive, and follicular-pattern results. This estimand was selected because the clinical question was whether the intraoperative result provided sufficient evidence to justify immediate surgical escalation. It is consistent with the clinical uncertainty represented by deferred and non-definitive findings in the primary studies [9-12].

The unit of analysis was the patient or nodule, according to the source study. The extracted variables were author, year, design, country and centre, Bethesda III sample size, surgery, intraoperative malignant results, intraoperative non-malignant or non-definitive results, final malignant and benign histology, true positives, false positives, false negatives, true negatives, and reported change in surgical management. The complete quantitative dataset used for the primary analysis is provided in Appendix 2. Supplementary Information.

### Risk of bias and applicability

Risk of bias and applicability concerns were assessed using QUADAS-2 across patient selection, index test, reference standard, and flow and timing [13]. Particular attention was paid to selective application of frozen section, exclusion of non-definitive results from accuracy calculations, incomplete separation of Bethesda III subgroups, and uncertainty about the independence of final histopathology from intraoperative findings.

### Statistical analysis

Study-level sensitivity and specificity were calculated from the reconstructed 2 × 2 tables as sensitivity = TP/(TP + FN) and specificity = TN/(TN + FP), respectively.

Random-effects synthesis was conducted on the logit scale. The within-study variances were specified as 1/*TP* + 1/*FN* for sensitivity and 1/*TN*+ 1/*FP* for specificity. A 0.5 continuity correction was used for zero cells. Cochran’s Q, τ^2^, and *I*^2^ were used as complementary measures of heterogeneity; Q was not treated as a substitute for a hierarchical diagnostic model.

All studies had zero false-positive results. Consequently, a full bivariate Reitsma model with freely estimated covariance was not identifiable. A pseudo-bivariate approximation was used, with sensitivity and specificity modelled on the logit scale and covariance set to zero. An approximate HSROC curve and confidence region were derived from the pooled logit estimates. The resulting AUC and diagnostic odds ratio were treated as model-based summaries rather than exact empirical measures [5, 14].

Sensitivity analyses included exclusion of Mao 2023 [9] and leave-one-out analyses. Forest plots were prepared for study-level sensitivity and specificity, and the resulting study-level display is shown in **Figure 2**. A comparative HSROC display was prepared for the analyses including and excluding Mao 2023 and is shown in **Figure 3**. Statistical calculations, reconstruction of diagnostic 2 × 2 tables, random-effects logit meta-analyses, sensitivity analyses, and approximate HSROC summaries were performed with the assistance of Microsoft Copilot Analyst Agent; the authors independently checked the extracted data, formulas, model assumptions, numerical outputs, and clinical interpretation, and retain full responsibility for the analyses and conclusions.

**Figure 1.**
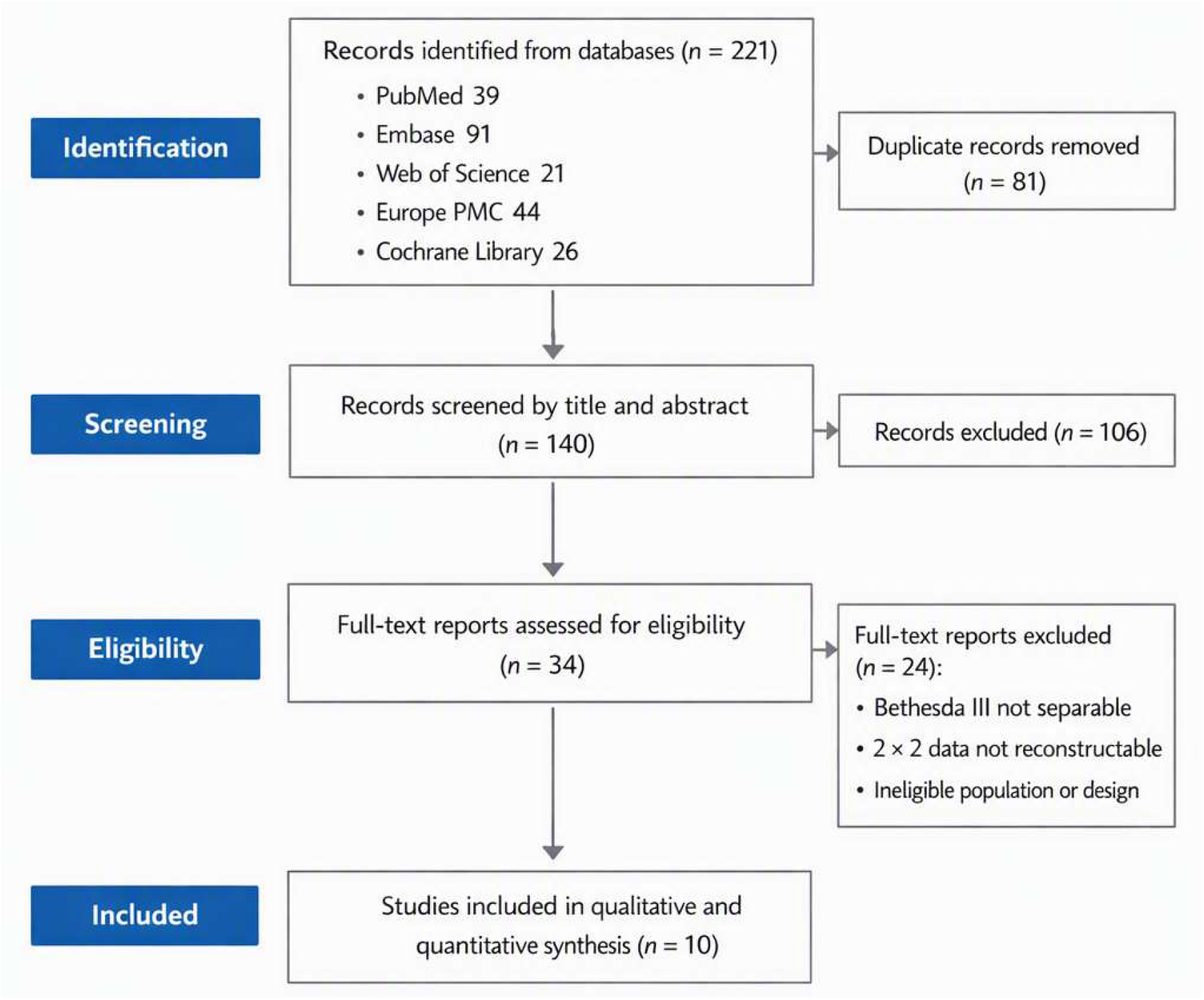
PRISMA-style study-selection flow diagram. Records identified, duplicates removed, records screened, full-text reports assessed, exclusions, and studies included in the qualitative and quantitative synthesis.

**Figure 2.**
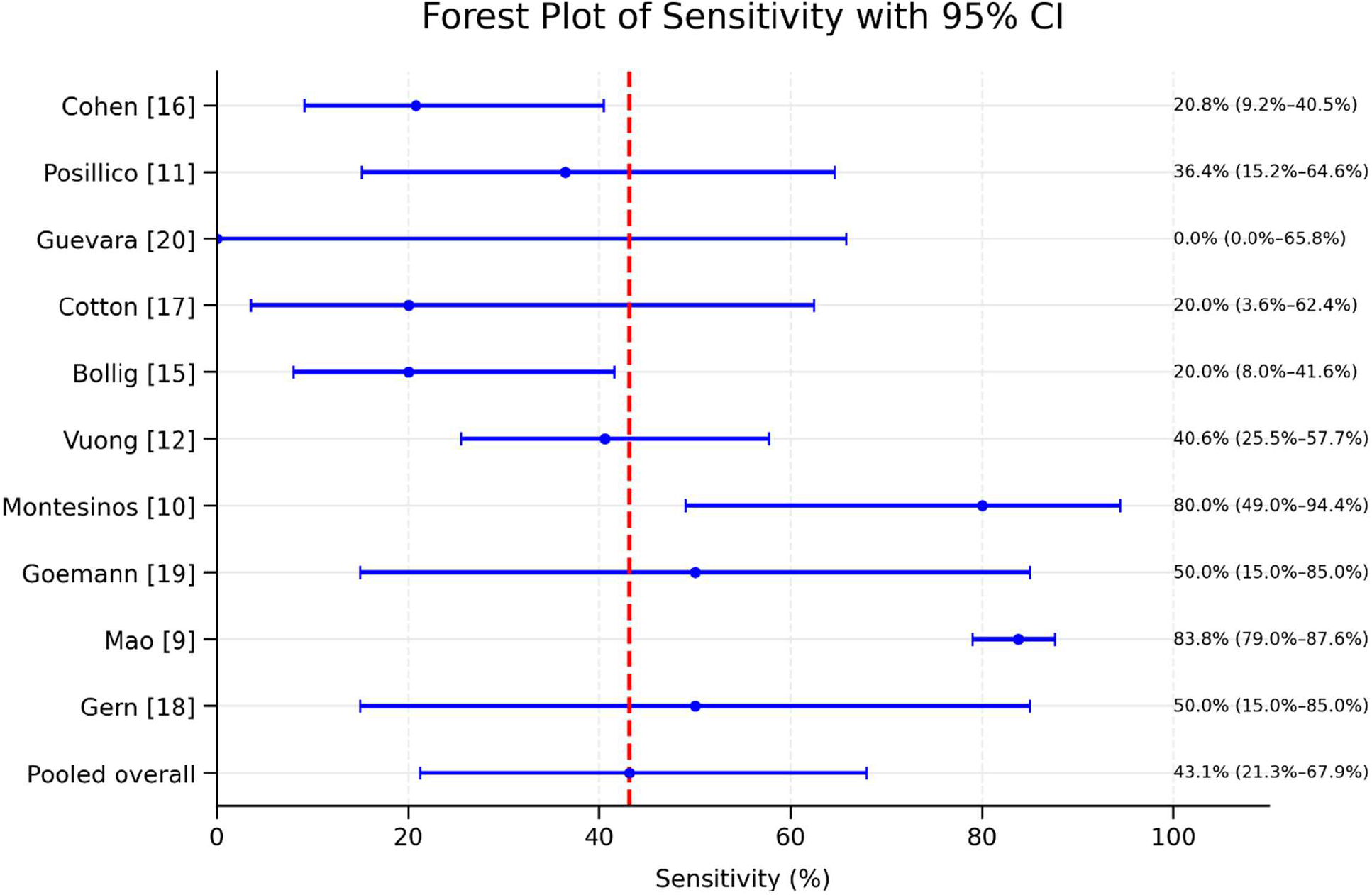
Text-rendered forest plot of study-level diagnostic accuracy. Study-level sensitivity and specificity estimates with Wilson 95% confidence intervals and pooled random-effects estimates. The figure is rendered from Table 3; pooled estimates are shown separately because they come from a random-effects logit model rather than from a simple average.

**Figure 3.**
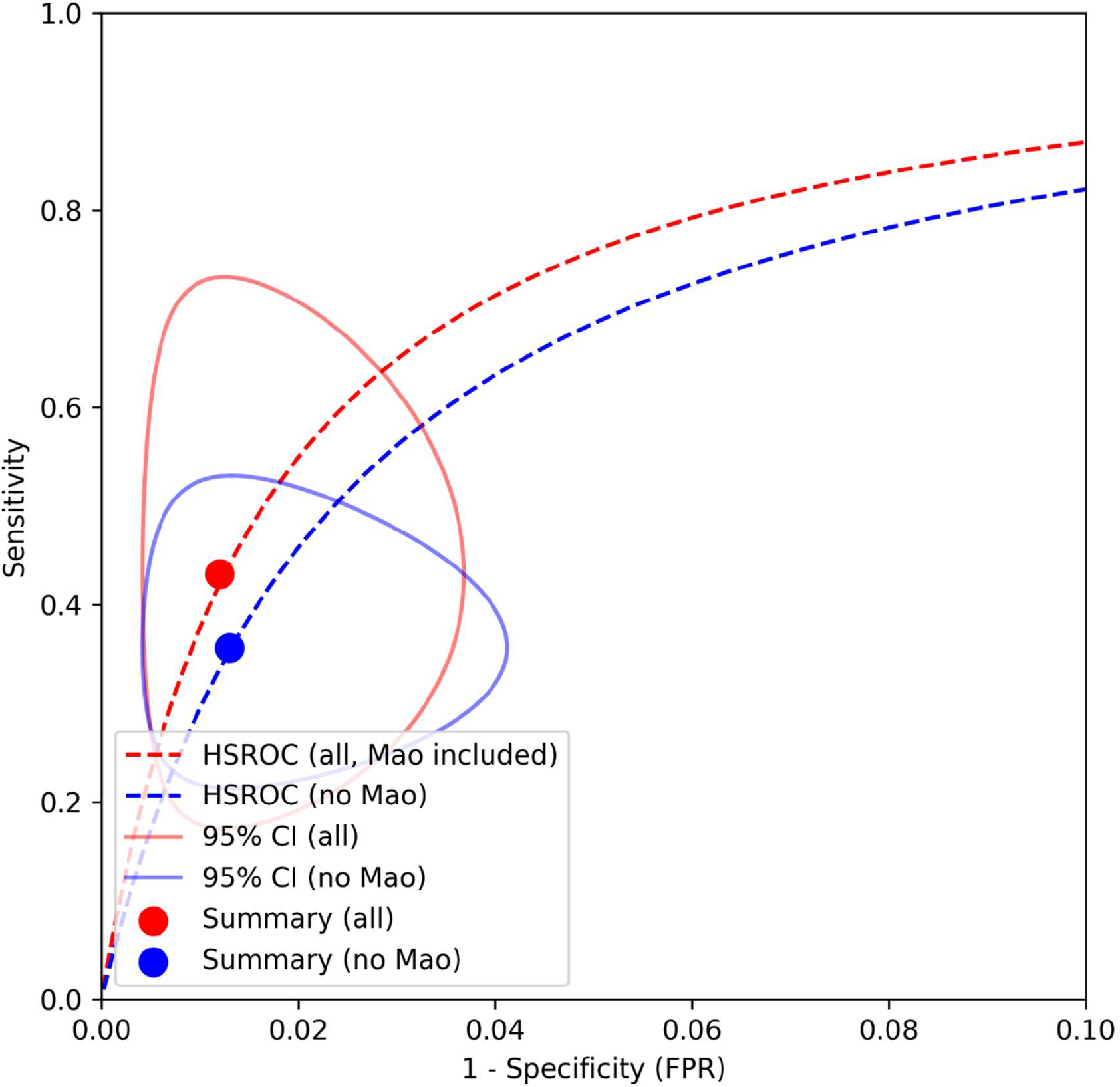
Comparative schematic HSROC display. False-positive rate is plotted on the horizontal axis and sensitivity on the vertical axis; summary points are shown for the analyses including and excluding Mao 2023. The curves remain in the high-specificity region, and their main difference is vertical displacement in sensitivity. The confidence region is therefore expected to be wider in the sensitivity direction and narrower in the false-positive direction. The apparent high overall discrimination is driven primarily by the near absence of false-positive results and should not be interpreted as evidence that a negative frozen-section result reliably excludes malignancy. Summary points: Including Mao 2023: sensitivity 0.431; specificity 0.988; FPR 0.012; AUC ≈ 0.89. Excluding Mao 2023: sensitivity 0.356; specificity 0.987; FPR 0.013; AUC ≈ 0.87

## RESULTS

### Study selection

The five database searches yielded 221 records: PubMed, 39; Embase, 91; Web of Science, 21; Europe PMC, 44; and Cochrane Library, 26. After 81 duplicates were removed, 140 unique records underwent title and abstract screening. Thirty-four full-text reports were assessed and 10 studies were included in the quantitative synthesis. Twenty-four full-text reports were excluded because Bethesda III was not separable, a 2 × 2 table could not be reconstructed, or the population or design was ineligible.

**Table 1.** Characteristics of studies included in the meta-analysis.

| Study (year) | Country | Bethesda III (n) | Intraoperative malignant | Final malignant | Final benign |
| --- | --- | --- | --- | --- | --- |
| Cohen 2015 [16] | United States | 170 | 5 | 24 | 146 |
| Posillico 2015 [11] | United States | 58 | 4 | 11 | 47 |
| Guevara 2015 [20] | France | 14 | 0 | 2 | 12 |
| Cotton 2016 [17] | United States | 45 | 1 | 5 | 40 |
| Bollig 2018 [15] | United States | 54 | 4 | 20 | 34 |
| Vuong 2020 [12] | United States | 314 | 13 | 32 | 282 |
| Montesinos 2021 [10] | Argentina | 21 | 8 | 10 | 11 |
| Goemann 2021 [19] | Brazil | 39 | 2 | 4 | 35 |
| Mao 2023 [9] | China | 342 | 232 | 277 | 65 |
| Gern 2024 [18] | Germany | 12 | 2 | 4 | 8 |

**Table 2.**
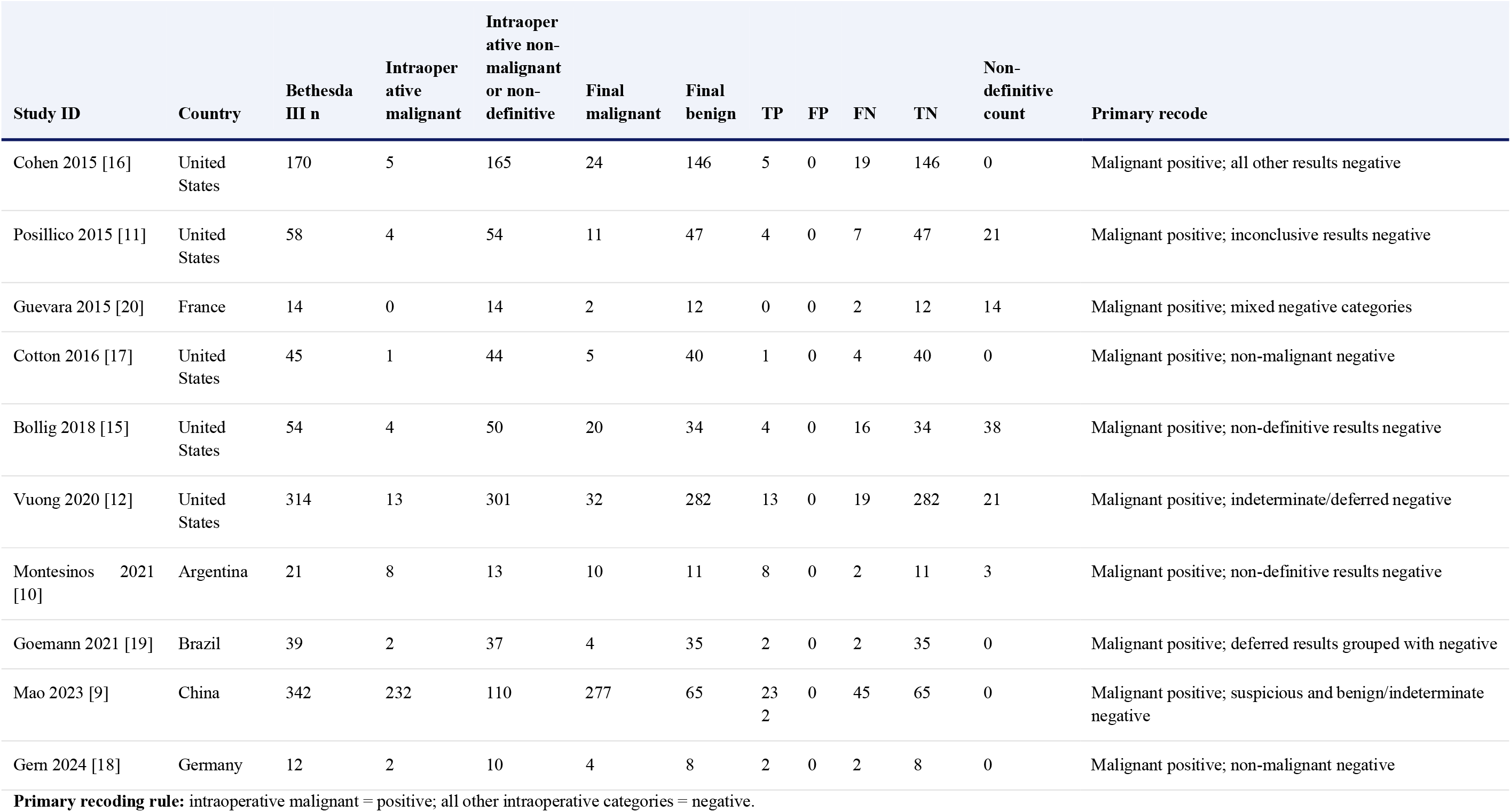
General study variables and reconstructed diagnostic data.

**Table 3.** Study-level sensitivity and specificity.

| Study | Sensitivity, % (95% CI) | Specificity, % (95% CI) | PPV, % | NPV, % |
| --- | --- | --- | --- | --- |
| Cohen 2015 [16] | 20.8 (9.2–40.5) | 100 (97.4–100) | 100 | 88.5 |
| Posillico 2015 [11] | 36.4 (15.2–64.6) | 100 (92.5–100) | 100 | 87.0 |
| Guevara 2015 [20] | 0.0 (0.0–65.8) | 100 (75.8–100) | Not estimable | 85.7 |
| Cotton 2016 [17] | 20.0 (3.6–62.4) | 100 (91.2–100) | 100 | 90.9 |
| Bollig 2018 [15] | 20.0 (8.0–41.6) | 100 (89.9–100) | 100 | 68.0 |
| Vuong 2020 [12] | 40.6 (25.5–57.7) | 100 (98.7–100) | 100 | 93.7 |
| Montesinos 2021 [10] | 80.0 (49.0–94.4) | 100 (74.1–100) | 100 | 84.6 |
| Goemann 2021 [19] | 50.0 (15.0–85.0) | 100 (90.1–100) | 100 | 94.6 |
| Mao 2023 [9] | 83.8 (79.0–87.6) | 100 (94.4–100) | 100 | 59.1 |
| Gern 2024 [18] | 50.0 (15.0–85.0) | 100 (67.6–100) | 100 | 80.0 |

The point estimates demonstrate the central pattern of the evidence: sensitivity varied substantially, whereas specificity was uniformly high. Because all false-positive cells were zero, PPV was 100% whenever at least one malignant intraoperative result was recorded.

The study-selection process is summarised in **Figure 1**.

### Study characteristics

The 10 included studies were published between 2015 and 2024 and came from the United States, France, Argentina, Brazil, China, and Germany. All were retrospective or retrospective cohort studies. The Bethesda III sample size ranged from 12 to 342, and the total number of analysed patients or nodules was 1,069. The malignancy proportion ranged from 29% to 81%, indicating substantial differences in the populations selected for surgery and frozen-section evaluation. Clinical, pathological, surgical, and diagnostic variables are combined in **Table 1**. The reconstructed diagnostic 2 × 2 data and surgical-management variables are shown in **Table 2 (Appendix 1)**.

### Study-level test accuracy

The study-level sensitivity ranged from 0% to 83.8%. Specificity was 100% in every study because no false-positive result was recorded. Study-level estimates and Wilson 95% confidence intervals are shown in **Table 3**; pooled estimates were calculated using the random-effects logit model.

### Pooled accuracy and heterogeneity

The pooled sensitivity was 43.1% (95% CI, 21.3–67.9) and the pooled specificity was 98.8% (95% CI, 97.0–99.5). The aggregate crude sensitivity, 271/389 or 69.7%, is not a pooled diagnostic estimate and is reported only to distinguish simple aggregation from random-effects synthesis. These estimates should not be interpreted without considering spectrum and selection bias; their clinical interpretation is summarised in **Table 4**.

**Table 4.** Clinical interpretation of study-level diagnostic accuracy estimates.

| Finding | Observed result | Clinical interpretation |
| --- | --- | --- |
| Sensitivity | 0.0% to 83.8% | Frozen section showed widely variable ability to detect malignancy across studies. |
| Lower sensitivity estimates | Cohen 2015, 20.8%; Cotton 2016, 20.0%; Bollig 2018, 20.0% | In these studies, frozen section identified only a minority of malignant nodules. |
| Intermediate sensitivity estimates | Posillico 2015, 36.4%; Vuong 2020, 40.6%; Goemann 2021, 50.0%; Gern 2024, 50.0% | The test detected approximately one third to one half of malignant cases. |
| Higher sensitivity estimates | Montesinos 2021, 80.0%; Mao 2023, 83.8% | These studies showed the highest detection rates; Mao 2023 was influential in the pooled analysis. |
| Specificity | 100% in every study | No false-positive results were recorded, so a malignant frozen-section result was consistently confirmed by final histopathology. |
| Positive predictive value | 100% when estimable; not estimable in Guevara 2015 | A positive frozen-section diagnosis was highly reliable for confirming malignancy. |
| Negative predictive value | 59.1% to 94.6% | A non-malignant or non-definitive frozen-section result did not consistently exclude malignancy. |
| Overall implication | High specificity and PPV, but variable sensitivity and inconsistent NPV | Frozen section is best interpreted as a rule-in test rather than a rule-out test in Bethesda III thyroid nodules. |

Sensitivity heterogeneity was extreme in the primary analysis: Q = 83.45 with 9 degrees of freedom, I^2^ >90%, and τ^2^ = 2.15. This very high heterogeneity indicates that the pooled sensitivity was strongly influenced by between-study variability and by the influential Mao 2023 study. After excluding Mao 2023, heterogeneity decreased to a moderate level (Q = 12.44 with 8 degrees of freedom, I^2^ = 36%, and τ^2^ = 0.25), indicating acceptable consistency between studies and a more robust pooled sensitivity estimate. Specificity showed negligible between-study variance. The heterogeneity pattern is consistent with substantial variation in case selection, pathology practice, and handling of non-definitive intraoperative categories; the pooled sensitivity heterogeneity statistics are summarised in **Table 5** [9, 11, 12, 15, 16].

**Table 5.**
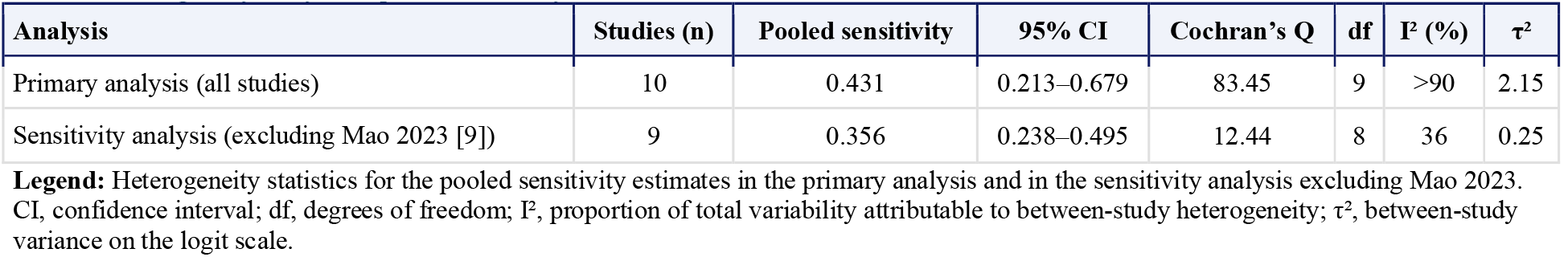
Heterogeneity analysis of pooled sensitivity.

| Analysis | Studies (n) | Pooled sensitivity | 95% CI | Cochran's Q | df | I<sup>2</sup> (%) | $\tau^2$ |
| --- | --- | --- | --- | --- | --- | --- | --- |
| Primary analysis (all studies) | 10 | 0.431 | 0.213–0.679 | 83.45 | 9 | >90 | 2.15 |
| Sensitivity analysis (excluding Mao 2023 [9]) | 9 | 0.356 | 0.238–0.495 | 12.44 | 8 | 36 | 0.25 |

**Legend:** Heterogeneity statistics for the pooled sensitivity estimates in the primary analysis and in the sensitivity analysis excluding Mao 2023. CI, confidence interval; df, degrees of freedom; I<sup>2</sup>, proportion of total variability attributable to between-study heterogeneity; $\tau^2$ , between-study variance on the logit scale.

A precision-driven required information size analysis indicated that the current evidence base, comprising 1,069 Bethesda III nodules and 389 malignant cases, exceeded the estimated number of malignant cases required to achieve a 95% confidence interval half-width of ±10% around the pooled sensitivity estimate. Using this clinically relevant precision threshold, no additional malignant cases or studies would be required to meet this precision target under either the high-heterogeneity scenario (τ^2^ = 2.15) or the reduced-heterogeneity scenario after excluding Mao 2023 (τ^2^ = 0.25). More stringent precision targets would require additional information, particularly for a ±5% half-width under the high-heterogeneity scenario. The required information size sensitivity analysis is summarised in **Table 6**.

**Table 6.** Required information size sensitivity analysis.

| Precision target | High heterogeneity scenario ($\tau^2 = 2.15$) | | | Reduced heterogeneity scenario ($\tau^2 = 0.25$) | | |
| --- | --- | --- | --- | --- | --- | --- |
|  | Required malignant cases | Additional malignant cases | Additional studies | Required malignant cases | Additional malignant cases | Additional studies |
| $\pm 10\%$ | 297 | 0 | 0 | 118 | 0 | 0 |
| $\pm 7.5\%$ | 528 | 139 | 4 | 209 | 0 | 0 |
| $\pm 5\%$ | 1,187 | 798 | 21 | 471 | 82 | 3 |

**Legend:** Required information size estimates for pooled sensitivity are shown under two assumptions: the primary high-heterogeneity model ( $\tau^2 = 2.15$ ) and the reduced-heterogeneity model after excluding Mao 2023 ( $\tau^2 = 0.25$ ). The current evidence base includes 389 malignant cases; therefore, the $\pm 10\%$ precision target is already met in both scenarios. The number of additional studies is approximate and was estimated from the mean malignant-case contribution of the included studies.

The study-level forest plot is shown in **Figure 2**. It provides a visual summary of the two main diagnostic patterns observed across studies. First, sensitivity estimates were widely dispersed, ranging from no intraoperative detection of malignancy in some cohorts to high sensitivity in the influential Mao 2023 study [9]. This dispersion indicates substantial between-study variability in the ability of frozen section to identify malignant Bethesda III nodules. Second, specificity estimates were tightly clustered at or near 100%, reflecting the absence of false-positive frozen-section results in the included studies. The confidence intervals were wider in smaller cohorts, particularly for sensitivity, whereas larger studies contributed more precise estimates. Overall, the forest plot supports the central interpretation of the meta-analysis: frozen section has a consistently strong rule-in value when the intraoperative result is malignant, but its variable sensitivity limits its value as a rule-out test when the result is benign, deferred, indeterminate, or otherwise non-definitive.

### Influence and sensitivity analyses

Excluding Mao 2023 reduced pooled sensitivity to 35.6% (95% CI, 23.8–49.5) and reduced sensitivity heterogeneity to Q = 12.44 with 8 degrees of freedom, *p* ≈ 0.13, τ^2^=0.^2^49, and *I*^2^ ≈ 36%. Specificity remained 98.7% (95% CI, 96.7–99.5), with negligible between-study variance.

The leave-one-out analysis showed little change when any study other than Mao 2023 was removed. Removal of Mao produced the major shift in sensitivity and heterogeneity. The finding supports treating Mao 2023 as an influential study in a prespecified sensitivity analysis rather than simply deleting it from the primary analysis.

### HSROC and ROC interpretation

The approximate HSROC analysis yielded a pooled diagnostic odds ratio of approximately 60 including Mao 2023 and approximately 41 excluding Mao 2023. The corresponding model-based AUC estimates were approximately 0.89 and 0.87, respectively. These are approximate summaries because the absence of false positives prevented estimation of a fully parameterised bivariate model. The comparative HSROC display is shown in **Figure 3**.

The coordinate system is false-positive rate on the horizontal axis and sensitivity on the vertical axis. All individual studies had a false-positive rate of zero and therefore lie on the vertical axis; the summary points are slightly to the right because the continuity correction was applied in the pooled model.

An HSROC curve is a hierarchical summary receiver operating characteristic curve used in diagnostic meta-analysis to summarise the trade-off between sensitivity and specificity across studies. Each study contributes a pair of values: sensitivity on the vertical axis and false-positive rate, calculated as 1 − specificity, on the horizontal axis. Points closer to the upper-left corner indicate better diagnostic performance, because they combine high sensitivity with a low false-positive rate. In this review, however, all studies had zero false-positive results, so the HSROC pattern is driven mainly by the very high specificity. Therefore, the high model-based AUC should not be interpreted as evidence that frozen section reliably excludes malignancy. Instead, the HSROC should be read as showing a test with strong rule-in value when positive, but limited rule-out value when the intraoperative result is negative or non-definitive.

### Risk of bias and applicability

No study had low risk of bias across all four QUADAS-2 domains. Six studies had at least one high-risk domain: Posillico 2015, Cotton 2016, Bollig 2018, Montesinos 2021, Mao 2023, and Gern 2024 [9-11, 15, 17, 18]. The remaining studies were judged to have unclear overall risk, mainly because the independence of final histopathology from the intraoperative interpretation was not adequately reported. The domain-level QUADAS-2 assessment is summarised in **Table 7**.

**Table 7.** QUADAS-2 summary.

| Domain | Low risk | Unclear risk | High risk | Main concern |
| --- | --- | --- | --- | --- |
| Patient selection | 2 | 3 | 5 | Selective frozen-section use and surgically enriched cohorts |
| Index test | 6 | 4 | 0 | Incomplete reporting of interpretation and thresholds |
| Reference standard | 0 | 10 | 0 | Appropriate histology, but independence or blinding unclear |
| Flow and timing | 7 | 2 | 1 | Exclusions or selective handling of non-definitive results |

Applicability concerns were high in patient selection for five studies, unclear in two, and low in three. Applicability of the reference standard was low concern in all studies. The main threat to clinical transportability was that the analysed cohorts consisted of patients selected for surgery and, in several studies, selected for frozen-section examination rather than all Bethesda III nodules.

## DISCUSSION

This systematic review and meta-analysis shows that intraoperative frozen section in Bethesda III thyroid nodules has a markedly asymmetric diagnostic profile: sensitivity is limited and heterogeneous, whereas specificity is consistently very high. The pooled specificity of 98.8% and the absence of false-positive results across the included studies indicate that a clearly malignant frozen-section result is unlikely to be false positive in the reported settings [9-12, 15-20].

The clinical limitation is the low sensitivity. The pooled estimate of 43.1% means that a substantial proportion of malignant nodules were not classified as malignant intraoperatively. After excluding Mao 2023, sensitivity fell to 35.6%, while heterogeneity decreased markedly. Thus, the more stable signal across the remaining studies is that frozen section detects only a minority to approximately one third of malignancies in this setting. A benign, deferred, indeterminate, suspicious, or follicular-pattern result should therefore not be interpreted as evidence that malignancy has been excluded [10-12, 15-20].

The influence analysis is clinically and methodologically important. Mao 2023 contributed 342 of the 1,069 analysed Bethesda III cases and reported an 81% malignancy prevalence in this subgroup [9]. The cohort was restricted to patients with available FNA, frozen-section, and final-pathology data, creating a strongly selected population. Its high sensitivity shifted the pooled estimate upward and accounted for much of the between-study heterogeneity. The study should therefore be retained in the primary analysis but reported prominently in sensitivity analyses rather than silently excluded.

The HSROC analysis provides a useful visual summary, but it requires careful interpretation. The approximate AUC values of 0.89 including Mao 2023 and 0.87 excluding Mao 2023 do not mean that the test has uniformly excellent clinical accuracy. Because every study had zero false-positive results, the ROC profile is dominated by specificity, while sensitivity remains uncertain and variable. The near-vertical appearance of the curves reflects a test that is strong for confirming malignancy when positive but weak for excluding malignancy when negative. Sparse-data diagnostic meta-analysis requires precisely this caution because continuity corrections and unstable covariance estimates can influence pooled ROC summaries [5, 14].

### Limitations of the study

All included studies were retrospective, and most were single-centre studies with small samples and non-uniform application of frozen section [9-12, 15-20]. Five studies had high risk of bias in patient selection, and no study had low risk of bias in all QUADAS-2 domains. These features limit the extent to which the estimates represent consecutive Bethesda III patients undergoing thyroid surgery [9-11, 13, 17, 18].

The large variation in malignancy prevalence, selective use of the index test, and inconsistent handling of non-definitive results may have distorted both sensitivity and specificity. Bollig 2018 illustrates this problem: 38 of 54 FLUS cases had non-definitive frozen-section results, and the original accuracy calculations were restricted to definitive results [15]. In the present review, non-malignant and non-definitive categories were recoded as negative to answer a clinically relevant question, but this recoding is an analytical decision and may not match the original estimand of every study.

The zero false-positive rate prevented estimation of a fully identified bivariate model with freely estimated covariance. The pseudo-bivariate HSROC, pooled DOR, and approximate AUC should therefore be regarded as model-based summaries under sparse-data constraints, not as definitive empirical ROC measures [5, 14]. The small number of studies also precluded reliable meta-regression, formal assessment of publication bias, and robust subgroup analysis. The certainty of evidence is low or very low because of risk of bias, inconsistency, and imprecision [21, 22].

The available evidence appears sufficient for clinically relevant precision thresholds. The current dataset includes 389 malignant cases, exceeding the estimated requirement for a ±10% confidence-interval half-width around the pooled sensitivity estimate. However, additional studies would be needed to achieve narrower precision targets, particularly ±7.5% or ±5%. Therefore, the main limitation of the evidence base is less the overall number of malignant cases than the substantial between-study heterogeneity, selective case inclusion, and risk of bias.

### Practical applicability

The findings are most applicable to patients already selected for thyroid surgery for Bethesda III nodules and treated in centres with intraoperative pathology expertise. In this context, a clearly malignant frozen-section result may support consideration of completion surgery during the same operation, provided that the decision is consistent with tumour extent, imaging, patient preferences, and the surgeon’s and pathologist’s judgment [3, 4].

The findings do not support routine frozen section for every Bethesda III nodule. A non-malignant or non-definitive result should not be used as the sole reason to avoid definitive surgery, restrict the extent of an otherwise indicated operation, or reassure the patient that cancer has been excluded. Frozen section is better understood as a selective rule-in tool: it may confirm malignancy when positive, but it cannot safely rule out malignancy when negative [4]. The practical value of the test is therefore greatest when a positive result would produce an immediate, clinically justified change in operative management. When the result would not alter the planned operation, or when the patient is not otherwise selected for surgery, the limited sensitivity and the risk of non-definitive findings reduce the rationale for routine use.

## CONCLUSIONS

Intraoperative frozen section in Bethesda III thyroid nodules has excellent specificity but limited and highly variable sensitivity. The test is useful mainly for confirming malignancy when the intraoperative diagnosis is clearly malignant. It is not a reliable rule-out test, and a benign, suspicious, deferred, or indeterminate result should not be used alone to exclude cancer or determine the extent of surgery. The available evidence supports selective rather than routine use, with interpretation integrated into the wider clinical, radiological, pathological, and patient-preference context.

## Data Availability

All data produced in the present work are contained in the manuscript

## DECLARATIONS

### Data availability

The complete quantitative dataset, search strategies, database yields, and selection flow are provided in Appendix 2. Supplementary Information. QUADAS-2 assessment and analysis files are stored in the project workspace.

### Author contributions

José Luis Pardal-Refoyo contributed to all stages of the study, including conceptualisation, protocol development, methodology, eligibility criteria, search planning, study selection, data extraction, reconstruction of diagnostic 2 × 2 tables, statistical analysis, interpretation of results, risk-of-bias assessment, drafting of the manuscript, critical revision, and approval of the final version. Ignacio Zapatero Sánchez contributed to study planning, bibliographic searching, and final review and approval of the manuscript.

## Appendix 1

## Appendix 2 Supplementary Information

## Diagnostic performance of intraoperative frozen section in Bethesda III thyroid nodules

**Last search date:** July 7, 2026.

### 1. Search strategies and database yields

The search combined three conceptual blocks:

- thyroid nodule or thyroid neoplasm;
- intraoperative frozen section or equivalent intraoperative diagnostic terms;
- Bethesda III or equivalent AUS/FLUS terminology.

No restrictions by publication date, language, or study design were documented. The results below correspond to records retrieved before deduplication.

| Database | Platform | Records retrieved | Search date |
| --- | --- | --- | --- |
| PubMed | National Library of Medicine | 39 | July 7, 2026 |
| Embase | Elsevier | 91 | July 7, 2026 |
| Web of Science | Web of Science Core Collection | 21 | July 7, 2026 |
| Europe PMC | Europe PMC | 44 | July 7, 2026 |
| Cochrane Library | Cochrane Library | 26 | July 7, 2026 |
| **Total** |  | **221** | **July 7, 2026** |

#### 1.1 PubMed

**(“T****hyroid** **N****eoplasms****”[M****esh****] OR “T****hyroid** **N****odule****”[M****esh****] OR** **thyroid nodule**[**tiab****] OR** **thyroid nodul**[**tiab****]) AND (“F****rozen** **S****ections****”[M****esh****] OR** **frozen section****\*[****tiab****] OR** **intraoperative consultation**[**tiab****] OR** **intraoperative diagnosis**[**tiab****]) AND (B****ethesda**[**tiab****] OR “B****ethesda** **III”[****tiab****] OR “B****ethesda** **3”[****tiab****] OR AUS[****tiab****] OR FLUS[****tiab****] OR** **atypia of undetermined significance**[**tiab****] OR** **follicular lesion of undetermined significance**[**tiab**])

**Records retrieved:** 39.

**Search date:** July 7, 2026.

#### 1.2 Embase

**(‘****thyroid tumor****’/****exp** **OR ‘****thyroid nodule****’/****exp** **OR ‘****thyroid nodule****’:****ti**,**ab**,**kw** **OR ((****thyroid** **NEAR/2** **nodul**):**ti**,**ab**,**kw****)) AND (‘****frozen section****’/****exp** **OR ‘****frozen section****\*’:****ti**,**ab**,**kw** **OR ‘****intraoperative consultation****’:****ti**,**ab**,**kw** **OR ‘****intraoperative diagnosis****’:****ti**,**ab**,**kw** **OR ((****intraoperative** **NEAR/2 (****consultation** **OR** **diagnosis**)):**ti**,**ab**,**kw****)) AND (****bethesda**: **ti**,**ab**,**kw** **OR ‘****bethesda iii****’:****ti**,**ab**,**kw** **OR ‘****bethesda** **3’:****ti**,**ab**,**kw** **OR** **aus**: **ti**,**ab**,**kw** **OR** **flus**: **ti**,**ab**,**kw** **OR ‘****atypia of undetermined significance****’:****ti**,**ab**,**kw** **OR ‘****follicular lesion of undetermined significance****’:****ti**,**ab**,**kw** **OR ((****atypia** **NEAR/3** **undetermined** **NEAR/3** **significance**):**ti**,**ab**,**kw**))

**Records retrieved:** 91.

**Search date:** July 7, 2026.

#### 1.3 Web of Science Core Collection

**TS= ((“****thyroid nodule****” OR (****thyroid** **NEAR/2** **nodul****) OR “****thyroid neoplasm****”)) AND TS=(“****frozen section****” OR “****intraoperative consultation****” OR “****intraoperative diagnosis****” OR (****intraoperative** **NEAR/2 (****consultation** **OR** **diagnosis****))) AND TS=(B****ethesda** **OR “B****ethesda** **III” OR “B****ethesda** **3” OR AUS OR FLUS OR “****atypia of undetermined significance****” OR “****follicular lesion of undetermined significance****” OR (****atypia** **NEAR/3** **undetermined** **NEAR/3** **significance**))

**Records retrieved:** 21.

**Search date:** July 7, 2026.

#### 1.4 Europe PMC

**(TITLE_ABS:****thyroid** **AND TITLE_ABS:****nodule****\*) AND (TITLE_ABS:”****frozen section****\*” OR TITLE_ABS:****intraoperative** **) AND (TITLE_ABS:B****ethesda** **OR TITLE_ABS:AUS OR TITLE_ABS:FLUS)**

**Records retrieved:** 44.

**Search date:** July 7, 2026.

#### 1.5 Cochrane Library

**#1 (B****ethesda** **OR “B****ethesda** **III” OR “B****ethesda** **3” OR AUS OR FLUS OR “****atypia of undetermined significance****” OR “****follicular lesion of undetermined significance****”) AND #2 (****frozen****\* NEXT** **section****\* OR** **intraoperative**)

**Records retrieved:** 26.

**Search date:** July 7, 2026.

### 2 Selection and deduplication flow

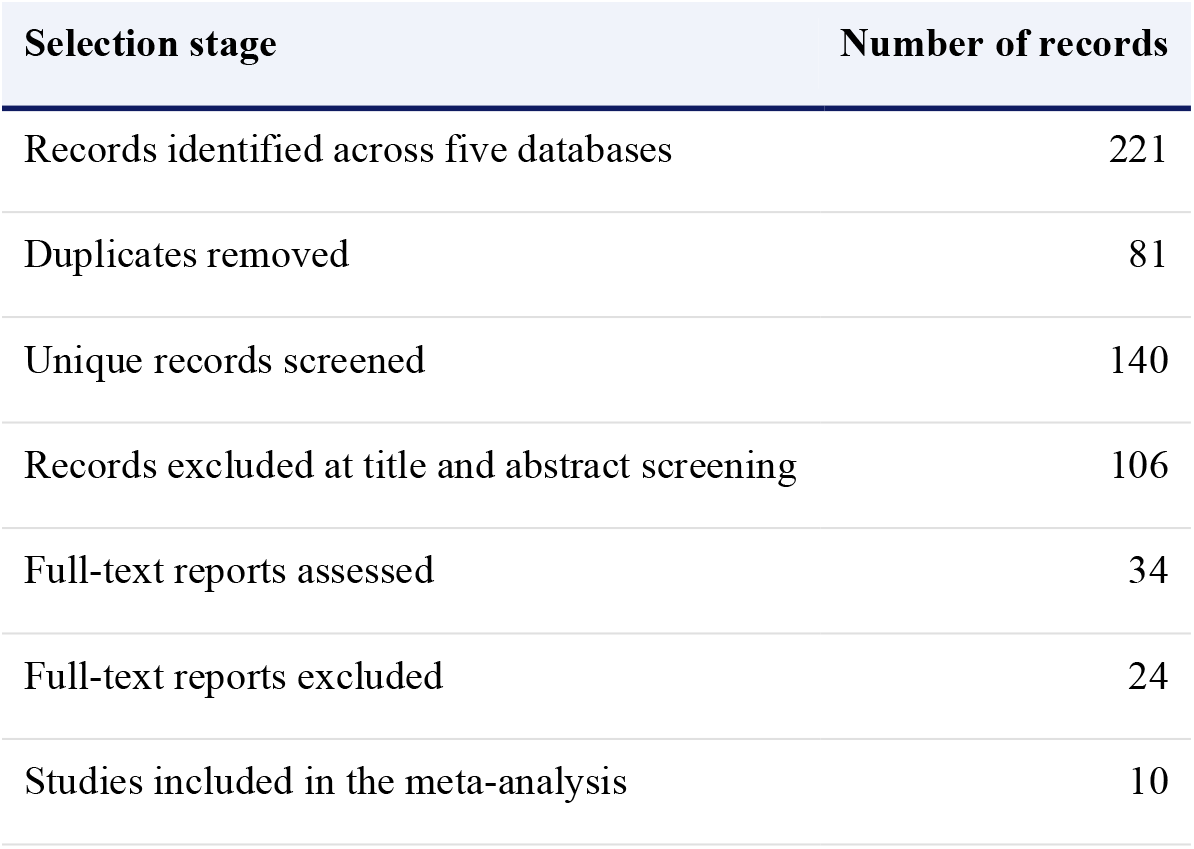

2. Selection and deduplication flow
| Selection stage | Number of records |
| --- | --- |
| Records identified across five databases | 221 |
| Duplicates removed | 81 |
| Unique records screened | 140 |
| Records excluded at title and abstract screening | 106 |
| Full-text reports assessed | 34 |
| Full-text reports excluded | 24 |
| Studies included in the meta-analysis | 10 |

### 3. Reproducibility note

The search strategies are transcribed from the project search record. For exact future replication, the database platform, local search time, complete export files, platform-specific syntax, and any interface filters should also be archived. The final search date for all five databases was July 7, 2026.

